# Evaluation of a new, community-based screening program to detect hearing loss in adult childhood cancer survivors in Switzerland – Findings from the HEAR study

**DOI:** 10.1101/2025.08.28.25334426

**Authors:** Philippa Jörger, Carina Nigg, Christina Schindera, Maša Žarković, Grit Sommer, Martin Kompis, Annika Frahsa, Nicolas Waespe, Marc Ansari, Claudia E. Kuehni

## Abstract

2.

**Purpose:** Childhood cancer survivors have an increased risk of long-term health complications, including treatment-related hearing loss. Although early detection is important, many adult survivors do not attend hearing screenings in clinical centers because visits can be logistically or emotionally burdensome. The HEAR study tested an alternative, community-based audiological screening option delivered in hearing aid shops in Switzerland. We evaluated its effectiveness, including clinical outcomes and survivor engagement, and developed a plan for potential implementation in clinical practice.

**Methods:** We invited childhood cancer survivors (CCS) registered in the Childhood Cancer Registry and diagnosed with cancer before age 21 years to a free pure-tone audiogram at hearing aid shops across Switzerland. Participants completed a baseline questionnaire before the hearing test, and two follow-up questionnaires evaluating feasibility and user experience. We gathered qualitative insights through semi structured interviews with participants and hearing aid shop employees, and group discussions with healthcare professionals. Interviews were analyzed using thematic analysis, and group discussions using template analysis. We evaluated the program according to the RE-AIM framework, incorporating both quantitative and qualitative data.

**Results:** Of 1604 invited CCS, 476 (30%) consented and 319 (20%) completed audiometric testing. The program identified clinically relevant hearing loss in 71 participants (22%) using the SIOP-Boston ototoxicity scale. Following the screening, five participants acquired hearing aids. Both CCS participants and clinicians were open to this alternative screening option and provided predominantly positive feedback. Together with clinicians, we developed an implementation plan that outlines how this screening option could be integrated into follow-up care.

**Conclusion:** This simple and accessible community-based screening option could complement existing follow-up care, particularly for CCS who are no longer engaged in structured follow-up care.

## 3. Introduction

For many survivors of cancer during childhood, care does not end with cure. Survivors face the risk of treatment-related late effects that include hearing loss from ototoxic treatments such as cranial radiation and platinum chemotherapy, affecting around 10% of all survivors [1,2]. Because these effects can appear years after treatment end, lifelong, risk-based follow-up care is essential for early detection and intervention [3–6].

In North America and Europe, risk-adapted follow-up care for pediatric childhood cancer survivors (CCS) is usually provided at oncology centers where treatment occurred [7–9]. All recommended examinations are organized in one location with specialists included as needed (e.g., ear nose throat [ENT] specialist for hearing). When CCS become adults, they transition to adult follow-up care. During transition, pediatric oncologists provide a treatment summary with individualized follow-up recommendations. This summary guides future care, which may be coordinated by an adult oncologist or hematologist, a general practitioner (GP), or through a structured long-term follow-up (LTFU) program with a multidisciplinary setup [7–9]. In Switzerland, several centers have LTFU programs for risk-based follow-up care [10,11], but due to their high resource demands, only a limited number of CCS can be served.

Many adult CCS do not attend recommended follow-up care [12–16]. In Switzerland, only half [13] and in France, only one-third of adult CCS reported attending follow-up care [12]. Factors that reduce participation in follow-up care include limited awareness of possible late effects [14,17], long travel distances for examinations [14,18,19], financial burdens related to medical visits [20,21], and the emotional impact of experiencing cancer and fear of adverse findings [22,23].

To increase participation in follow-up, the HEAR study tested a new screening option for CCS based in hearing aid shops [24]. Hearing aid shops in Switzerland offer hearing tests for free to anyone. This option allows survivors to receive high quality audiological testing in a community-based setting, outside medical facilities, which could reduce organizational effort, cost, and psychological burden. We determined feasibility, effectiveness, limitations, and implementation requirements of this screening option using the Reach, Effectiveness, Adoption, Implementation and Maintenance (RE-AIM) framework [25,26].

## 4. Methods

### 4.1 Study design

The HEAR study, a health service research project launched in 2021 at the University of Bern, Switzerland, evaluated a community-based hearing screening program for CCS. The study protocol has been published [24]. Participants were invited for audiometric testing at hearing aid shops across Switzerland. We conducted a multimethod evaluation of the program using questionnaires, interviews, and group discussions with participants and stakeholders that included hearing aid shop employees and clinicians in (pediatric) oncology, follow-up care, and ENT specialization. The RE-AIM framework guided the study design and evaluation [25,26]. As this pilot study assessed feasibility rather than long-term integration, we combined the implementation and maintenance domains. Supplementary Table 1 shows the RE-AIM domains contextualized to our study.

The Ethics Committee of the Canton of Bern approved the HEAR study (2021-01624). Participants provided informed written consent for study participation.

### 4.1 Study population

CCS eligible for the HEAR study were identified through the Childhood Cancer Registry (ChCR), Switzerland’s national cancer registry for children diagnosed before age 21 [27,28]. We invited individuals aged ≥18 years, German or French-speaking, and potentially at risk for treatment-related hearing loss due to chemotherapy or radiation to head, neck, or spine [1,29–32]. We distinguished a high risk group treated with cranial radiation ≥30 Gray or cisplatin/carboplatin, the standard risk group consisted of those who received any other chemotherapy or cranial radiation <30 Gray, or any neck/spine radiation.

### 4.2 The HEAR study screening program

All eligible CCS (N=1604) were invited by postal mail, with up to two reminders for nonresponders (Figure 1). Consenting participants completed an online baseline questionnaire and independently scheduled an appointment at a hearing aid shop of their choice, all belonging to one nationwide company.

**Figure 1:**
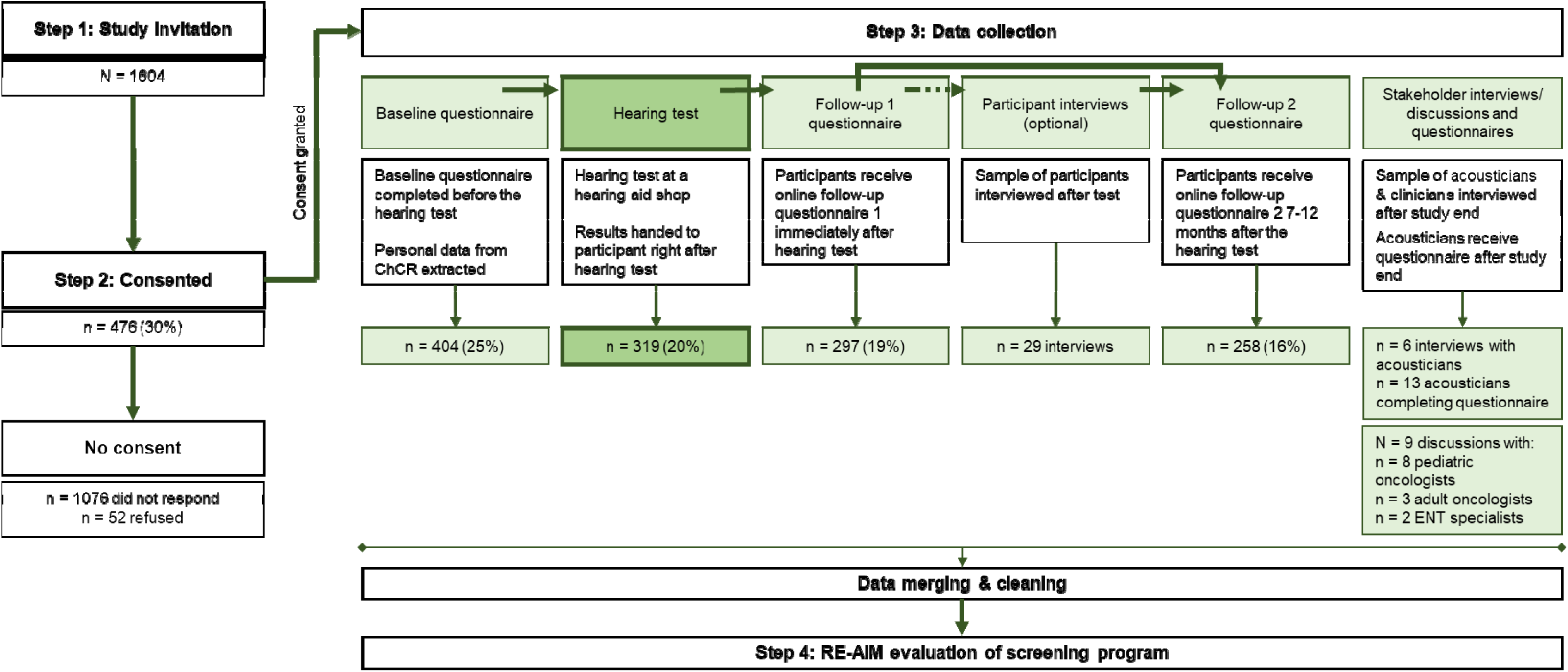
Study procedures and response overview. Abbreviations: ChCR, Childhood Cancer Registry. n, number. Hearing tests were completed between July 2022 and July 2023.

At the hearing aid shop, certified hearing aid acousticians (hearing aid technicians) performed bilateral pure tone audiometry (125 to 8000 Hz), the standard method for hearing loss screening [33]. If hearing loss exceeded 25 dB, bone conduction was measured (250 to 4000 Hz). Participants received their results and acousticians advised them to consult their physician if hearing problems or unclear findings emerged. For confidentiality reasons, acousticians were not informed of the medical history of participants.

### 4.3 Data collection

#### 4.3.1 CCS participants’ characteristics and audiograms

The ChCR provided clinical data of all eligible CCS on sex, cancer diagnosis, age at diagnosis, current age, treatment, correspondence language, and address. We derived the Swiss Neighborhood Index of Socioeconomic Position (Swiss-SEP) [34] and degree of urbanization (urban, periurban, rural) [35] based on address. We classified Swiss-SEP into tertiles (low, middle, high) [34]. Hearing aid shops submitted the audiogram results to the research team in a coded format. More details on data sources have been published previously [24].

#### 4.3.2 Questionnaires

The baseline questionnaire before the hearing test assessed sociodemographic characteristics, reasons for participating, and self-reported hearing. Participants received a follow-up questionnaire immediately after the assessment, including questions about their experiences with the hearing test, and another one 7-12 months later, including questions about actions taken since the hearing test, and perceived facilitators and barriers (Figure 1). We collected and managed questionnaire data using the Research Electronic Data Capture tool (REDCap version 13.7.5, Vanderbilt University, Nashville, TN, USA, 2022) [36,37].

#### 4.3.3 Interviews and group discussions

We collected qualitative data through semistructured interviews and group discussions with participants and key stakeholders. We interviewed 29 study participants and five acousticians who performed a hearing test for the study. The latter informed an online questionnaire we sent to a larger sample of acousticians. We further invited clinicians from different centers in Switzerland, including eight pediatric oncologists, three adult oncologists/hematologists, and two ENT specialists to one-on-one and small group discussions (1–3 per session) to explore how such a screening option could be integrated into existing follow-up care practices.

### 4.4 Data analysis

#### 4.4.1 Quantitative data analysis

We analyzed quantitative data using frequencies and descriptive statistics to summarize key measures across the RE-AIM dimensions. To compare groups, we used chi-square tests and independent t-tests. We performed all analyses in STATA version 16 (Stata Corporation, Austin, TX, USA, 2023) and RStudio (R Core Team, Vienna, AUT, 2022).

We assessed hearing loss using the International Society of Pediatric Oncology (SIOP) Boston Ototoxicity Scale, which is used for early detection of hearing loss in CCS exposed to ototoxic treatments [38]. We considered sensorineural hearing loss and the grading of the more affected ear. Severity was graded as none (grade 0), mild (grade 1), moderate (grade 2), or severe hearing loss (≥ grade 3) [38]. Clinically relevant hearing loss was defined as grade ≥ 2 [39].

#### 4.4.2 Qualitative data analysis

We recorded all interviews and group discussions. We transcribed interviews in Word before importing into MAXQDA 2022 (VERBI Software, Berlin, DE, 2021) for analysis.

During group discussions, one researcher systematically took minutes, which a second researcher supplemented and verified.

Interviews with study participants were analyzed using reflexive thematic analysis [40]. Detailed methodology along with the results of the study participant interviews are published elsewhere [41]. We applied principles of template analysis to structure and analyze qualitative input from clinicians and hearing aid shop employees [42,43].

## 5. Results

### 5.1 Reach

Among 1604 invited CCS, 476 (30%) consented and 319 (20%) completed a hearing test (in the following referred to as participants) (Figure 1). Follow-up questionnaire 1 was returned by 297 CCS (19%), and follow-up questionnaire 2 by 258 (16%). Half (53%) of participants were female, median age at study was 33 years (interquartile range [IQR]: 26– 41), age at diagnosis 8 years (IQR: 4–14), and median time since diagnosis 26 years (IQR: 18–34) (Table 1). Dominant diagnoses were leukemias (n=104, 33%) and lymphomas (n=84, 26%). Overall, 83 (26%) were categorized as high risk.

**Table 1:** Characteristics of participants completing a hearing test. (N=319)

|  | Completed hearing test (N=319) | Did not complete hearing test (N=1285) | Total (N=1604) | P-value<sup>a</sup> |
| --- | --- | --- | --- | --- |
|  | n (%) | n (%) | n (%) |  |
| **Sex (female)** | 168 (53%) | 518 (40%) | 686 (43%) | <0.01 |
| **Age at study, median (IQR)** | 33 (26-41) | 29 (23-38) | 30 (23-39) | <0.01 |
| 18-24 | 66 (21%) | 417 (32%) | 483 (30%) |  |
| 25-34 | 111 (35%) | 445 (35%) | 556 (35%) |  |
| 35-44 | 96 (30%) | 315 (25%) | 411 (26%) |  |
| 45-59 | 46 (14%) | 108 (8%) | 154 (10%) |  |
| **Years since diagnosis, median (IQR)** | 26 (18-34) | 21 (13-31) | 22 (14-31) | <0.01 |
| **Age at diagnosis, median (IQR)** | 8 (4-14) | 10 (4-14) | 10 (4-14) | 0.22 |
| **ICCC-3 Diagnosis** |  |  |  | 0.17 |
| Leukemias | 104 (33%) | 458 (36%) | 562 (35%) |  |
| Lymphomas | 84 (26%) | 308 (24%) | 392 (24%) |  |
| CNS tumors | 17 (5%) | 112 (9%) | 129 (8%) |  |
| Other | 114 (36%) | 407 (32%) | 521 (32%) |  |
| **Risk group** |  |  |  | 0.85 |
| Standard risk group | 236 (74%) | 944 (74%) | 1180 (74%) |  |
| High risk group | 83 (26%) | 341 (27%) | 424 (26%) |  |
| **Still in regular follow-up care<sup>c,d</sup>** |  |  |  | NA |
| Yes | 120 (38%) | NA | NA |  |
| No | 198 (62%) | NA | NA |  |
| **Language** |  |  |  | 0.07 |
| French | 95 (30%) | 318 (25%) | 413 (26%) |  |
| German | 224 (70%) | 967 (75%) | 1191 (74%) |  |
| **Urbanicity<sup>b,a</sup>** |  |  |  | 0.03 |
| Urban | 190 (60 %) | 811 (63 %) | 1001 (63 %) |  |
| Periurban | 64 (20 %) | 288 (22 %) | 352 (22 %) |  |
| Rural | 64 (20 %) | 183 (14 %) | 247 (15 %) |  |
| **Swiss-SEP, median (IQR)<sup>b</sup>** | 61 (53-68) | 59 (51-66) | 59 (51-67) | 0.01 |
| Low | 86 (27%) | 447 (35%) | 533 (33 %) | 0.02 |
| Middle | 116 (36%) | 436 (34%) | 552 (34 %) |  |
| High | 117 (37%) | 401 (31%) | 518 (32 %) |  |
| **Place of birth of parents<sup>c,d</sup>** |  |  |  | NA |
| Both parents born in Switzerland | 225 (72%) | NA |  |  |
| One parent born abroad | 49 (16%) | NA | NA |  |
| Both parents born abroad | 40 (13%) | NA | NA |  |
| **Highest level of education<sup>c,d</sup>** |  |  |  | NA |
| Primary education | 15 (5%) | NA | NA |  |
| Secondary education | 147 (46%) | NA | NA |  |

|  | **Completed**<br>**hearing test**<br>**(N=319)** | **Did not complete**<br>**hearing test**<br>**(N=1285)** | **Total**<br>**(N=1604)** | **P-value<sup>a</sup>** |
| --- | --- | --- | --- | --- |
|  | n (%) | n (%) | n (%) |  |
| Tertiary education | 155 (49%) | NA | NA |  |

Table with column percentages.
Abbreviations: ICCC-3, international classification of childhood cancer, edition 3. IQR, interquartile range. n, number. NA, not applicable.
<sup>a</sup> P-values calculated using chi-square statistics for categorical and t-test for continuous variables.
<sup>b</sup> Urbanicity and social assistance rate at municipal level: According to the Swiss Federal Office for Statistics; Typology and economic social welfare at municipality level [51,52].
<sup>c</sup> Only available for participants that completed the baseline questionnaire.
<sup>d</sup> Missings excluded.

Participants were older, more often female, and further away from diagnosis than nonparticipants. They were more often French-speaking, residents of rural areas and had a higher socioeconomic position measured by the Swiss-SEP index. The most common reasons to participate were contribution to science (n=370, 92%), and checking their hearing (n=223, 55%), followed by the test being part of their follow-up care (n=76, 19%) and the test being free of charge (n=54, 13%) (Supplementary Figure 1).

### 5.2 Effectiveness

We identified clinically relevant hearing loss in 71 (22%) participants (Figure 2). Of those, 34 (15%) participants were in the standard risk group and 37 (44%) in the high risk group.

**Figure 2:**
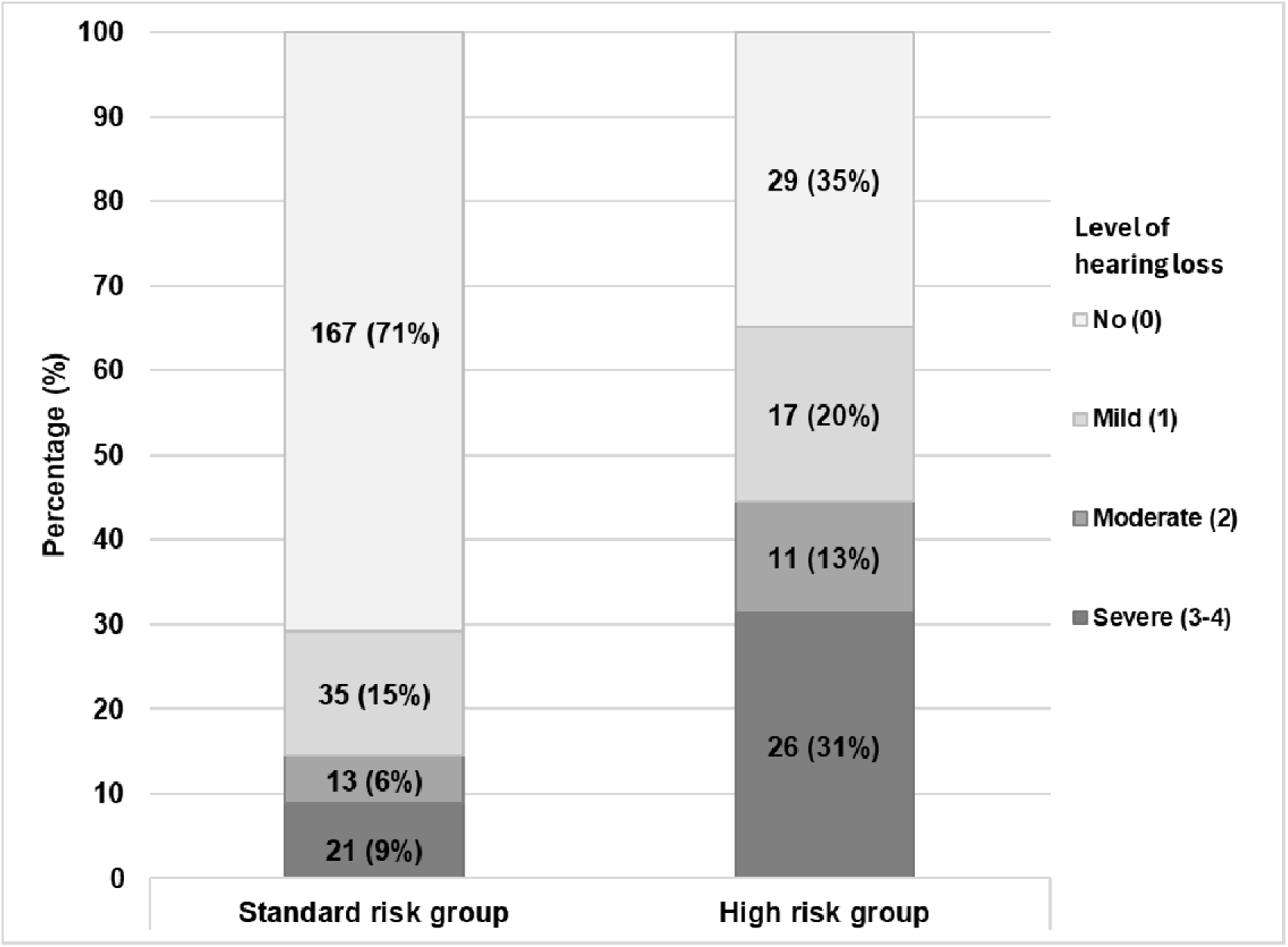
Prevalence and severity of hearing loss classified according to SIOP-Boston ototoxicity scale of participants completing a hearing test (N=319) stratified by risk group (high or standard risk group based on treatment exposure). Abbreviations: SIOP, International Society of Pediatric Oncology. Risk groups: High, treated with platinum agents (cisplatin/carboplatin) or cranial radiation .30 Gray. Standard, treated with any other chemotherapy or radiation to head, neck or spine with any dose. Hearing loss results from the high-risk group have been published previously [3].

Of 59 participants with clinically relevant hearing loss who completed follow-up questionnaire 2, 13 (22%) indicated they had been unaware of their hearing loss or that the degree had changed (Figure 3). Nineteen (32%) did not report hearing loss, possibly reflecting differences in result communication at hearing aid shops (Supplementary Table 2). Following the screening, 19 participants (32%) visited a physician and 5 of them (8%) acquired a new hearing aid.

**Figure 3:**
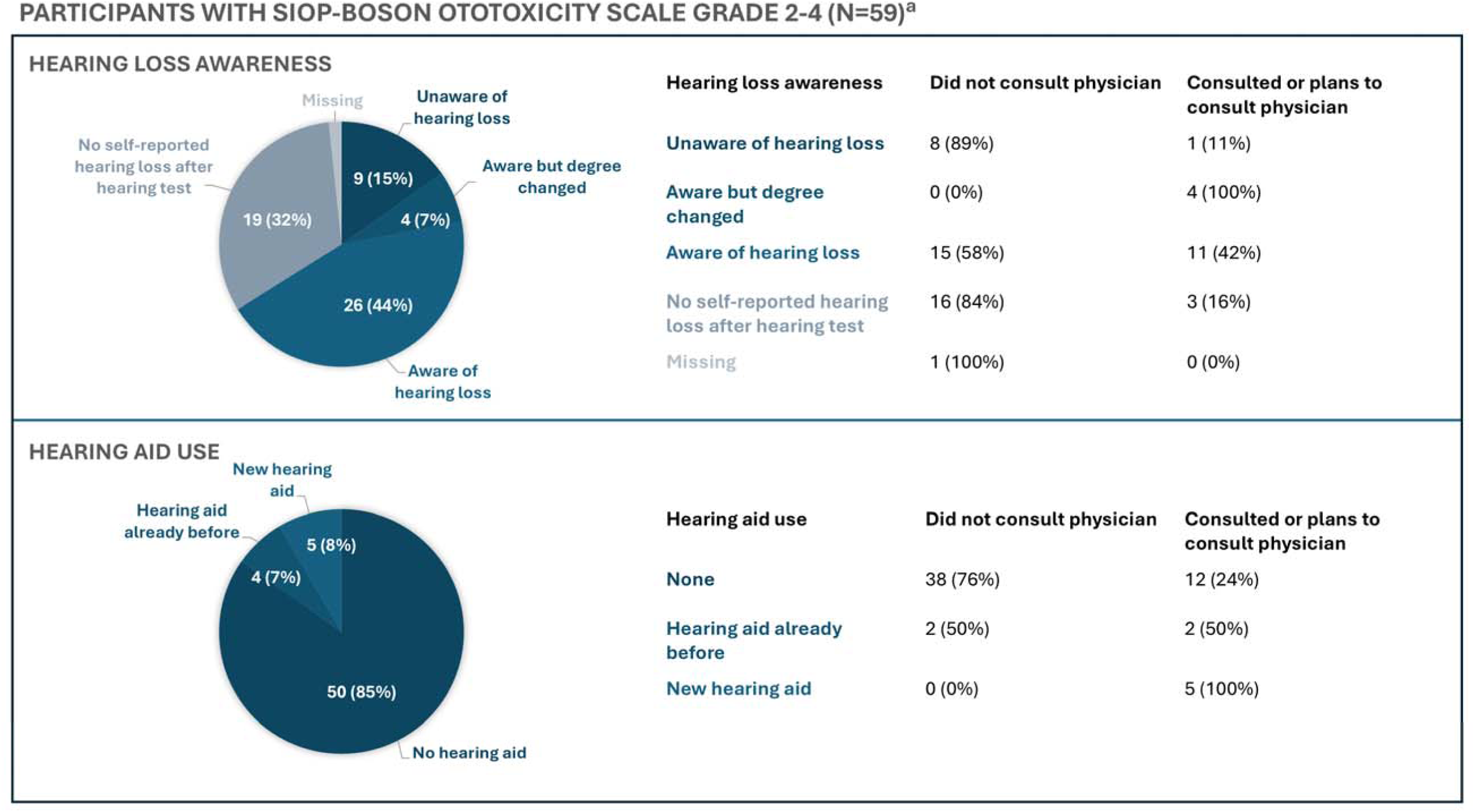
Hearing loss awareness and hearing aid use among participants with clinically relevant hearing loss (SIOP-Boston ototoxicity scale 2-4). Total and stratified by attended consultation with physicians after the hearing test. ^a^Participants with clinically relevant hearing loss according to SIOP Boston ototoxicity scale grade 2-4 who completed follow-up questionnaire 2.

### 5.3 Adoption

#### 5.3.1 CCS’ experiences

Of 296 participants to follow-up questionnaire one, 261 (88%) reported that their expectations of the hearing test were met (Supplementary Table 3). Nearly all participants (n=286, 97%) felt they had sufficient time to ask the acoustician questions, and 261 (88%) stated the results were clearly explained to them. Yet, 93 (32%) stated they did not receive or did not remember receiving information on how to proceed with the results, indicating need for improvement in post-screening guidance. Some participants reported in open questions that they expected more instructions on further screenings and managing hearing loss. Participants with clinically relevant hearing loss were more likely to prefer an immediate consultation with a physician about their results compared to those with normal hearing (41% vs. 17%, respectively).

Most participants traveled less than 30 minutes to a hearing aid shop (n=228, 77%), and it took less than 30 minutes to complete the test (n=167, 56%).

When asked about facilitators and barriers to audiometric screening across settings, participants rated hearing aid shops comparable to private practices or hospitals regarding professionalism, how seriously their concerns were taken, specialist empathy, and clarity of explanations (Figure 4). More participants found hearing aid shops easier to access, offering simpler appointment scheduling, quicker travel, more time to ask questions, and shorter visits in general.

**Figure 4:**
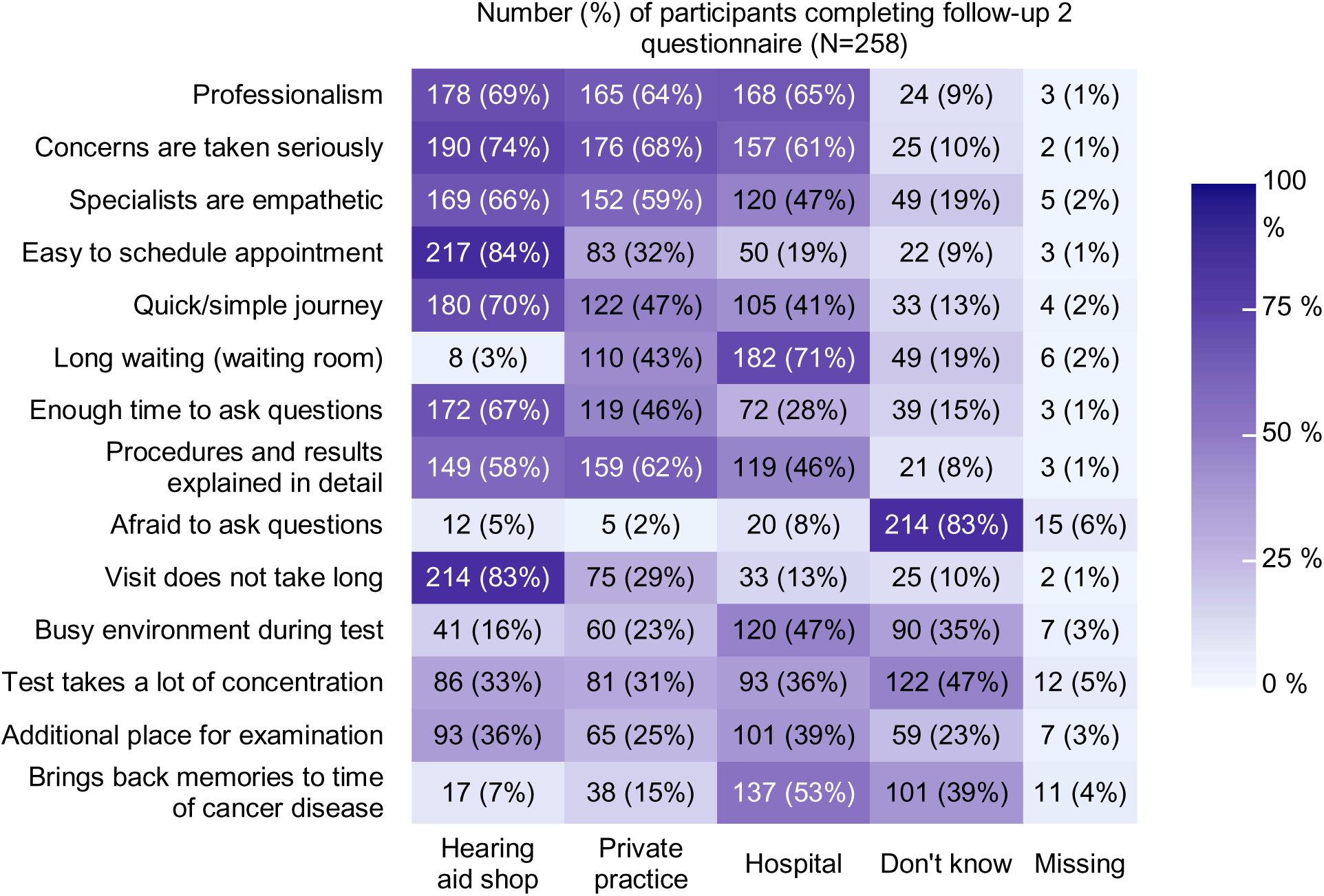
Heat map indicating proportions of participants reporting on facilitators and barriers for hearing screening in follow-up care: Comparing hearing aid shops, private practice and hospitals. Abbreviations: N, number. Higher intensity of color corresponds to a higher proportion.

Interviewed participants appreciated the program’s convenience and ease of integration into daily life. Some expressed reservations about the hearing test being an additional appointment to schedule, while structured follow-up care with central organization includes scheduling for them. Many valued the informal and welcoming atmosphere of hearing aid shops, others preferred a more medical environment where they could discuss results immediately with a physician. Detailed interview findings are published elsewhere [41].

#### 5.3.2 Hearing aid shop employee experiences

Challenges encountered on the providers’ side were study-specific and not expected in routine implementation. The lead coordinator from the hearing aid shops noted time consuming extraction of audiogram data from their database and staff turnover during the yearlong study, requiring study-related retraining of staff. The hearing aid provider initially opened 29 out of >200 stores for the study. After 11 invited CCS reported excessively long journeys to one of the selected shops, access was extended to additional shops. Ultimately, participants visited 55 different locations.

Acousticians reported positive experiences with participants. Among the 13 acousticians completing the questionnaire, none encountered any problems with study participants (data not shown). Acousticians perceived participants as less effortful and time intensive than usual customers (Supplementary Figure 2). While most felt well prepared to care for study participants (n=9), three acousticians felt the absence of medical background information that they could not assess due to confidentiality restrictions (Supplementary Figure 3). Were such screening routinely carried out, CCS would then be treated as regular customers and acousticians would not be challenged by confidentiality restrictions.

Interviewed acousticians expressed interest in more education on effects of ototoxic treatments to be able to better interpret results and respond to questions from CCS; this was echoed by one pediatric oncologist.

### 5.4 Implementation and maintenance

We summarized clinicians’ input on the screening option regarding target group, ways to inform CCS, implementation requirements, and further suggestions. This informed an implementation plan for integrating the screening option into routine follow-up care (Table 2).

**Table 2:**
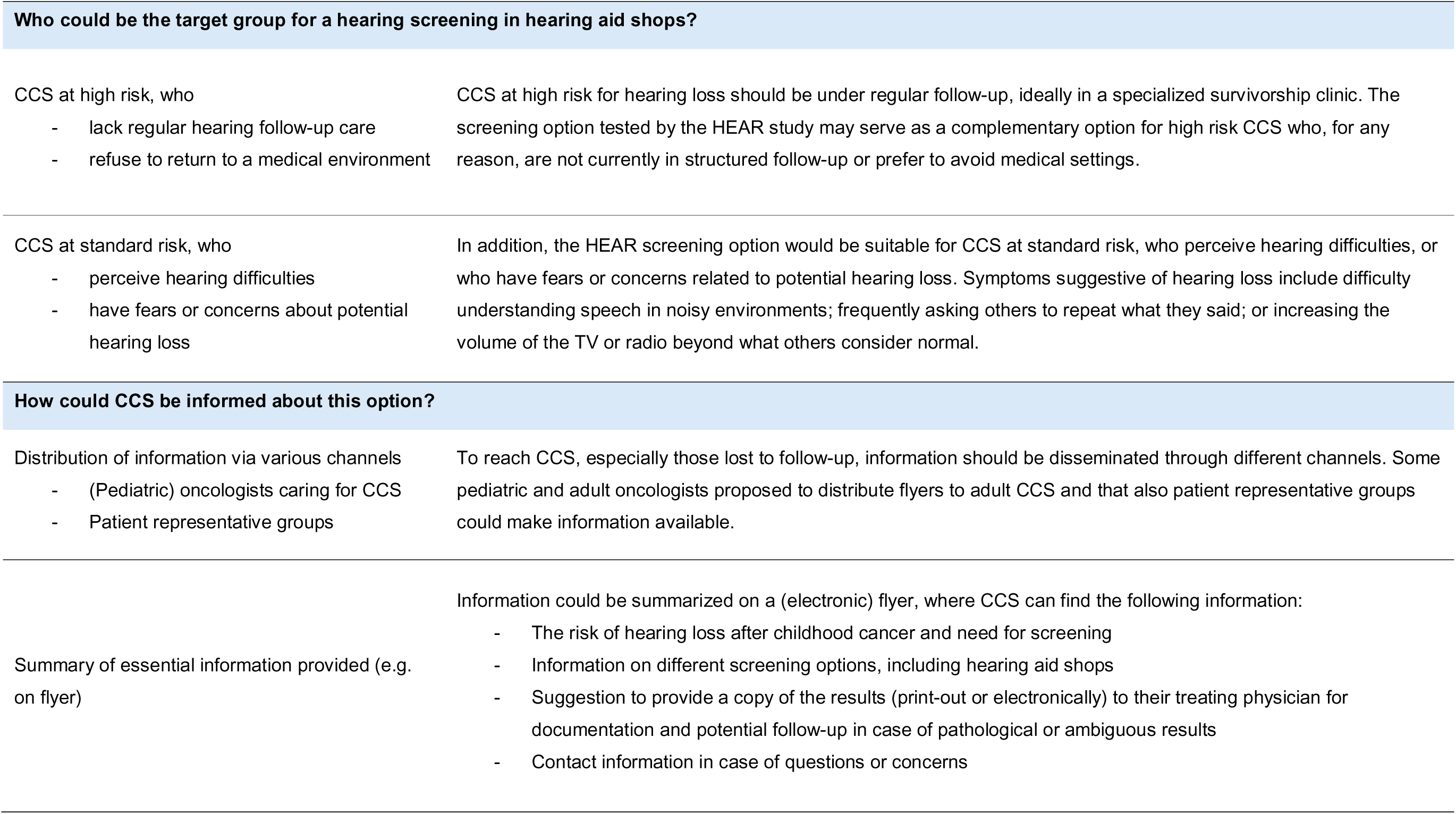
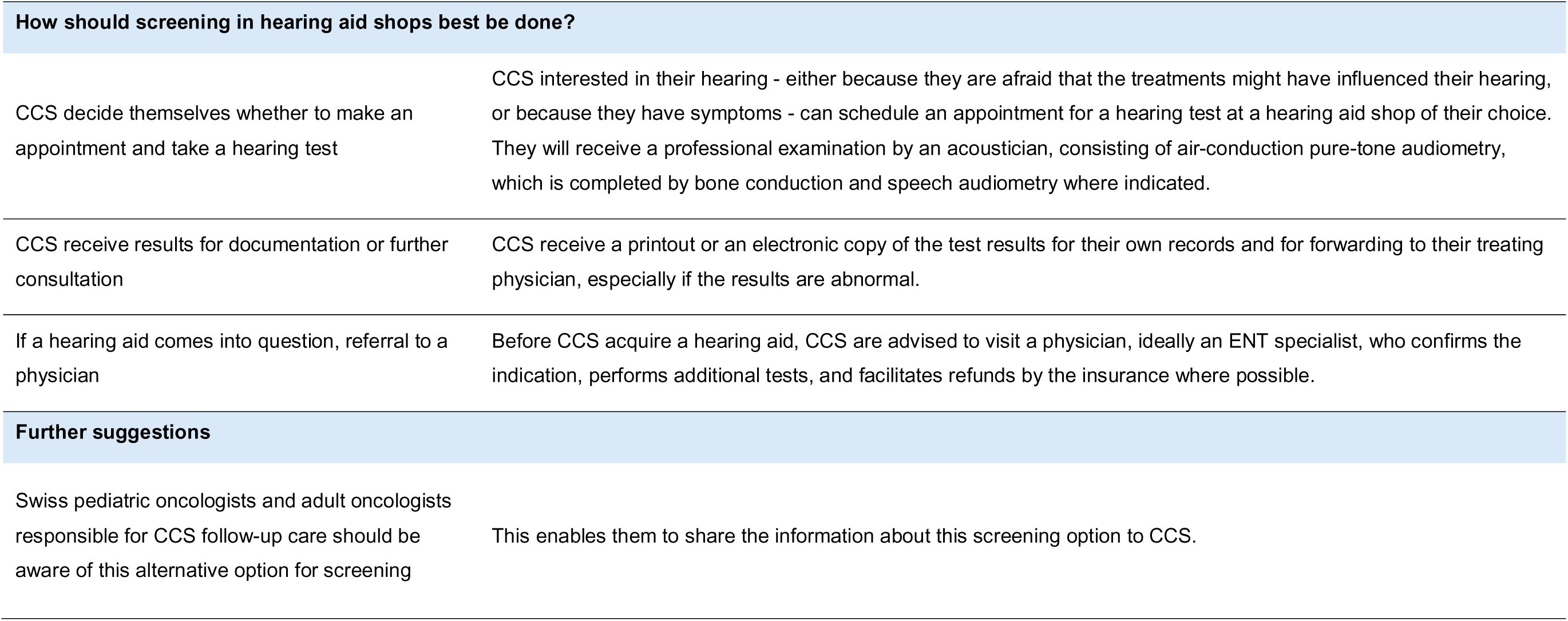
Implementation plan based on collected input from hearing aid shop employees, pediatric oncologists, adult cologists/hematologists involved in CCS’ follow-up care and ENT specialists.

| Who could be the target group for a hearing screening in hearing aid shops? |  |
| --- | --- |
| CCS at high risk, who<br>- lack regular hearing follow-up care<br>- refuse to return to a medical environment | CCS at high risk for hearing loss should be under regular follow-up, ideally in a specialized survivorship clinic. The screening option tested by the HEAR study may serve as a complementary option for high risk CCS who, for any reason, are not currently in structured follow-up or prefer to avoid medical settings. |
| CCS at standard risk, who<br>- perceive hearing difficulties<br>- have fears or concerns about potential hearing loss | In addition, the HEAR screening option would be suitable for CCS at standard risk, who perceive hearing difficulties, or who have fears or concerns related to potential hearing loss. Symptoms suggestive of hearing loss include difficulty understanding speech in noisy environments; frequently asking others to repeat what they said; or increasing the volume of the TV or radio beyond what others consider normal. |
| How could CCS be informed about this option? |  |
| Distribution of information via various channels<br>- (Pediatric) oncologists caring for CCS<br>- Patient representative groups | To reach CCS, especially those lost to follow-up, information should be disseminated through different channels. Some pediatric and adult oncologists proposed to distribute flyers to adult CCS and that also patient representative groups could make information available. |
| Summary of essential information provided (e.g. on flyer) | Information could be summarized on a (electronic) flyer, where CCS can find the following information:<br>- The risk of hearing loss after childhood cancer and need for screening<br>- Information on different screening options, including hearing aid shops<br>- Suggestion to provide a copy of the results (print-out or electronically) to their treating physician for documentation and potential follow-up in case of pathological or ambiguous results<br>- Contact information in case of questions or concerns |
| How should screening in hearing aid shops best be done? |  |
| CCS decide themselves whether to make an appointment and take a hearing test | CCS interested in their hearing - either because they are afraid that the treatments might have influenced their hearing, or because they have symptoms - can schedule an appointment for a hearing test at a hearing aid shop of their choice. They will receive a professional examination by an acoustician, consisting of air-conduction pure-tone audiometry, which is completed by bone conduction and speech audiometry where indicated. |
| CCS receive results for documentation or further consultation | CCS receive a printout or an electronic copy of the test results for their own records and for forwarding to their treating physician, especially if the results are abnormal. |
| If a hearing aid comes into question, referral to a physician | Before CCS acquire a hearing aid, CCS are advised to visit a physician, ideally an ENT specialist, who confirms the indication, performs additional tests, and facilitates refunds by the insurance where possible. |
| Further suggestions |  |
| Swiss pediatric oncologists and adult oncologists responsible for CCS follow-up care should be aware of this alternative option for screening | This enables them to share the information about this screening option to CCS. |

Clinicians were generally supportive, praising accessibility and flexibility of the screening option. They acknowledged that CCS can be tested outside a medical institution to reduce burden of already overbooked follow-up appointments.

Most agreed that the program is best suited for CCS who are not otherwise engaged in structured LTFU. For CCS in structured LTFU with established workflows external hearing tests were considered less valuable. While high risk CCS should generally be monitored within structured LTFU, most clinicians felt that the screening option could still be a valuable alternative for high risk individuals who are not currently in such care or who are reluctant to return to a clinical setting. In addition, some clinicians saw a benefit for CCS - regardless of risk level - who perceive symptoms of hearing loss or are concerned about their hearing. The alternative screening option could also be used by survivors of cancer in adulthood.

Clinicians suggested different methods to propagate this screening option. Some clinicians were open to distribute flyers to their patients, others recommended to ask patient organizations, or the Swiss Childhood Cancer Survivor Study [44] to inform CCS. One adult oncologist proposed to include information in the referral letter to the responsible GP or in the Survivorship Passport or Passport for Care [45,46]. Information to CCS should be carefully worded to prevent anxiety and include contact information in case of questions or concerns.

Clinicians recommended that all oncologists involved in follow-up care across Switzerland should be aware of the screening option so that they could provide information to interested survivors. Further, it was important to clinicians that any test results get back to the treating physician, ensuring integration into the broader follow-up care context and for documentation. Some expressed concerns about relying on CCS to return the hearing test results to physicians, questioning whether CCS would consistently share them. After the hearing test, CCS should get clear guidance on the next steps, especially if results are abnormal.

Few oncologists expressed concerns about hearing aid shops having a commercial interest in selling hearing aids. ENT specialists noted that existing mechanisms mitigate this risk: Acousticians typically refer individuals with a hearing problem to an ENT specialist for further tests. They evaluate the need for hearing aids and prescribe them, a requirement for insurance reimbursement. One ENT specialist highlighted that hearing aid providers have an interest in maintaining credibility, as poor practice could lead to negative recommendations from ENTs.

## 6. Discussion

### 6.1 Summary of main findings

This evaluation of a community-based hearing screening program demonstrated its feasibility and accessibility, and highlighted its potential to complement existing follow-up care, particularly in settings without centrally organized programs or when survivors do not attend regular follow-up. Study participants and stakeholders expressed general openness toward this screening option and provided predominantly positive feedback.

### 6.2 RE-AIM evaluation interpretation

Twenty percent of CCS completed a hearing test. This response is slightly lower than in a study using internet-based hearing test screening at home (29%) [47]. A reason for this could be that in contrast to internet-based testing at home, participants in our study had to schedule an appointment themselves and go there. Participating survivors were older than nonparticipants, which is probably a result of hearing loss becoming more prevalent with age [48] or because older participants may not have had a hearing test since undergoing treatment due to lack of organized follow-up care or limited knowledge about ototoxic late effects at that time. This aligns with our findings showing that high risk CCS treated longer ago were less aware about the risk of hearing loss [49]. Thus, older CCS and those not in follow-up care anymore may be an important target group for this screening option. Additionally, participants lived more often in rural areas, where the community-based option may be particularly appealing.

Based on the SIOP-Boston criteria, 15% of participants in the standard risk group and 44% in the high risk group showed clinically relevant hearing loss. The prevalence in the high risk group aligns with our previous findings in a different study population, suggesting that 36% of high risk CCS had hearing loss [50]. There is limited research on hearing loss among CCS not treated with platinum-based chemotherapy or cranial radiation. One U.S. study investigating a low risk group found impaired hearing in 23% of CCS [30]. However, that study employed more sensitive screening criteria. In our study, 22% of participants with clinically relevant hearing loss were unaware of their condition or noted a change in severity. Five individuals acquired a new hearing aid as a result of the screening, nearly doubling the number of hearing aid users in the sample. This highlights the effectiveness of the screening option to identify undetected hearing impairments. Importantly, all participants who obtained a new hearing aid consulted a physician, suggesting that the intended care pathway from screening to medical consultation and intervention works well.

The program was well accepted by participating CCS and acousticians. For CCS who prioritize convenience or feel apprehensive about medical settings, the community-based option appeared particularly suitable. The input from stakeholders guided the development of a feasible implementation plan that considered different follow-up care structures, communication pathways, and informational needs of CCS and providers.

### 6.3 Strengths and limitations

This study contributes to the field through piloting a community-based hearing screening program for CCS to complement existing follow-up care structures. Using the RE-AIM framework and a mixed-methods approach, we comprehensively evaluated the program’s feasibility, effectiveness, acceptability, and potential for sustainable implementation. We included the understudied population of CCS at standard risk for hearing loss.

While the program was designed as a low-threshold initiative, to ensure broad geographic accessibility across Switzerland, the hearing aid provider only opened a small proportion out of >200 shops, limiting access in some regions. In addition to physical accessibility, study-related ethical requirements also posed a barrier. The ethics committee in Switzerland has high standards: even for questionnaire and simple observational studies comprehensive and detailed patient information texts are required. While necessary for research, this would not apply if integrated in routine follow-up care, potentially increasing participation. Finally, since the screening was offered to CCS at a single time point, we could not assess its long-term usability in routine follow-up care. Additional barriers may still emerge during broader implementation.

### 6.4 Conclusion

This study examined a new hearing screening option designed to provide CCS with an alternative to screenings in medical settings. Its findings suggest that screening based in hearing aid shops is feasible and may be particularly well suited for CCS who do not attend follow-up care, value convenience, or feel reluctant to engage with medical institutions. We developed a pragmatic implementation plan that builds on existing follow-up care infrastructure and capacities of involved stakeholders enabling sustainable integration into long-term survivor care. The next step is to propagate this screening option in a real-world setting.

## Data Availability

The data that support the information of this manuscript were accessed on secured servers of the Institute of Social and Preventive Medicine at the University of Bern. Individual-level, fully anonymized, sensitive data can only be made available for researchers who fulfil the respective legal requirements. Requests of data from the Childhood Cancer Registry must be directed to the Childhood Cancer Registry of Switzerland (https://www.childhoodcancerregistry.ch).

## 9. Declarations

## 9.1 Acknowledgement

We thank the study team of the Childhood Cancer Research Group, Institute of Social and Preventive Medicine, University of Bern, the Swiss Childhood Cancer Survivor Study, and the team of the Swiss Childhood Cancer Registry. We thank Nina Hofer for her valuable work. We thank all childhood cancer survivors participating in the study and for the interesting discussions. We thank Jörg Beyer, Claudia Candreia, Lara Chavaz, Tamara Diesch-Furlanetto, Rhoikos Furtwängler, Fabienne Gumy Pause, Fatime Krasniqi, Sonja Lüer, Roby Mathew, Maria Otth, Eva Maria Tinner and Nicolas von der Weid for their valuable inputs. We thank the hearing aid provider and their employees for taking part in this study and providing audiometric screening to participants. We thank Christopher Ritter for editorial assistance.

## 9.2 Declarations

Authors PJ, CN, MŽ, GS, MK, AF and CK have no relevant financial or nonfinancial interests to disclose. NW reports a relationship with Swedish Orphan Biovitrum AB that includes advisory board membership, consulting, and travel reimbursement and a relationship with Novartis that includes advisory board membership. CS reports a relationship to Swedish Orphan Biovitrum AB that includes travel reimbursement. MA reports a relationship with Jazz Pharmaceutical and Novo Nordisk that includes travel reimbursement. None of these relationships has any association with the current study.

## References

1 Weiss A, Sommer G, Kasteler R, et al. Long-term auditory complications after childhood cancer: A report from the Swiss Childhood Cancer Survivor Study. Pediatr Blood Cancer. 2017;64:364–73. doi: 10.1002/pbc.26212

2 Robison LL, Hudson MM. Survivors of childhood and adolescent cancer: life-long risks and responsibilities. Nat Rev Cancer. 2014;14:61–70. doi: 10.1038/nrc3634

3 Winther JF, Kenborg L, Byrne J, et al. Childhood cancer survivor cohorts in Europe. Acta Oncol. 2015;54:655–68. doi: 10.3109/0284186X.2015.1008648

4 Michel G, Mulder RL, van der Pal HJH, et al. Evidence-based recommendations for the organization of long-term follow-up care for childhood and adolescent cancer survivors: a report from the PanCareSurFup Guidelines Working Group. J Cancer Surviv. 2019;13:759–72. doi: 10.1007/s11764-019-00795-5

5 International Guideline Harmonization Group. International Guideline Harmonization Group. 2024. https://www.ighg.org/ (accessed 1 February 2025)

6 Skinner R, Wallace WHB, Levitt G. Long-term follow-up of children treated for cancer: why is it necessary, by whom, where and how? Arch Dis Child. 2007;92:257–60. doi: 10.1136/adc.2006.095513

7 Guilcher GMT, Fitzgerald C, Pritchard S. A questionnaire based review of long-term follow-up programs for survivors of childhood cancer in Canada. Pediatr Blood Cancer. 2009;52:113–5. doi: 10.1002/pbc.21701

8 Eshelman-Kent D, Kinahan KE, Hobbie W, et al. Cancer survivorship practices, services, and delivery: a report from the Children’s Oncology Group (COG) nursing discipline, adolescent/young adult, and late effects committees. J Cancer Surviv. 2011;5:345–57. doi: 10.1007/s11764-011-0192-8

9 Essig S, Skinner R, von der Weid NX, et al. Follow-up programs for childhood cancer survivors in Europe: a questionnaire survey. PLoS One. 2012;7:e53201. doi: 10.1371/journal.pone.0053201

10 Babecoff S, Mermillod F, Marino D, et al. Long-term follow-up for childhood cancer survivors: the Geneva experience. Swiss Med Wkly. 2022;152:w30153. doi: 10.4414/SMW.2022.w30153

11 Tinner EM, Gumy Pause F, Diezi M, et al. Long-term follow-up after childhood cancer in Switzerland: a position statement from the pediatric Swiss LTFU working group. Bulletin suisse du cancer. 2019;39:212–5.

12 Dumas A, Milcent K, Bougas N, et al. Predictive factors of long-term follow-up attendance in very long-term childhood cancer survivors. Cancer. 2023;129:3476–89. doi: 10.1002/cncr.34944

13 Baenziger J, Roser K, Mader L, et al. Can the theory of planned behavior help explain attendance to follow-up care of childhood cancer survivors? Psychooncology. 2018;27:1501–8. doi: 10.1002/pon.4680

14 Ernst M, Brähler E, Faber J, et al. A Mixed-Methods Investigation of Medical Follow-Up in Long-Term Childhood Cancer Survivors: What Are the Reasons for Non-Attendance? Front Psychol. 2022;13:846671. doi: 10.3389/fpsyg.2022.846671

15 Rebholz CE, von der Weid NX, Michel G, et al. Follow-up care amongst long-term childhood cancer survivors: a report from the Swiss Childhood Cancer Survivor Study. Eur J Cancer. 2011;47:221–9. doi: 10.1016/j.ejca.2010.09.017

16 Michel G, Kuehni CE, Rebholz CE, et al. Can health beliefs help in explaining attendance to follow-up care? The Swiss childhood cancer survivor study. Psychooncology. 2011;20:1034–43. doi: 10.1002/pon.1823

17 Hendriks MJ, Harju E, Michel G. The unmet needs of childhood cancer survivors in long-term follow-up care: A qualitative study. Psychooncology. 2021;30:485–92. doi: 10.1002/pon.5593

18 Nathan PC, Agha M, Pole JD, et al. Predictors of attendance at specialized survivor clinics in a population-based cohort of adult survivors of childhood cancer. J Cancer Surviv. 2016;10:611–8. doi: 10.1007/s11764-016-0522-y

19 Michel G, Gianinazzi ME, Eiser C, et al. Preferences for long-term follow-up care in childhood cancer survivors. Eur J Cancer Care (Engl). 2016;25:1024–33. doi: 10.1111/ecc.12560

20 Cai J, Cheung YT, Hudson MM. Care Models and Barriers to Long-Term Follow-Up Care Among Childhood Cancer Survivors and Health Care Providers in Asia: A Literature Review. JCO Glob Oncol. 2024;10:e2300331. doi: 10.1200/GO.23.00331

21 Howard AF, Kazanjian A, Pritchard S, et al. Healthcare system barriers to long-term follow-up for adult survivors of childhood cancer in British Columbia, Canada: a qualitative study. J Cancer Surviv. 2018;12:277–90. doi: 10.1007/s11764-017-0667-3

22 Casillas J, Kahn KL, Doose M, et al. Transitioning childhood cancer survivors to adult-centered healthcare: insights from parents, adolescent, and young adult survivors. Psychooncology. 2010;19:982–90. doi: 10.1002/pon.1650

23 Rosenberg-Yunger ZRS, Klassen AF, Amin L, et al. Barriers and Facilitators of Transition from Pediatric to Adult Long-Term Follow-Up Care in Childhood Cancer Survivors. J Adolesc Young Adult Oncol. 2013;2:104–11. doi: 10.1089/jayao.2013.0003

24 Jörger P, Nigg C, Mader L, et al. A Health Service Research Study on a Low-Threshold Hearing Screening Program for Childhood Cancer Survivors in Switzerland: Protocol for the HEAR Study. JMIR Res Protoc. 2025;14:e63627. doi: 10.2196/63627

25 Glasgow RE, Vogt TM, Boles SM. Evaluating the public health impact of health promotion interventions: the RE-AIM framework. Am J Public Health. 1999;89:1322–7. doi: 10.2105/ajph.89.9.1322

26 Glasgow RE, Harden SM, Gaglio B, et al. RE-AIM Planning and Evaluation Framework: Adapting to New Science and Practice With a 20-Year Review. Front Public Health. 2019;7:64. doi: 10.3389/fpubh.2019.00064

27. Childhood Cancer Registry. Childhood Cancer Registry (ChCR). 2025. https://www.childhoodcancerregistry.ch/ (accessed 1 February 2025)

28 Michel G, von der Weid NX, Zwahlen M, et al. The Swiss Childhood Cancer Registry: rationale, organisation and results for the years 2001-2005. Swiss Med Wkly. 2007;137:502–9. doi: 10.4414/smw.2007.11875

29 Clemens E, van den Heuvel-Eibrink MM, Mulder RL, et al. Recommendations for ototoxicity surveillance for childhood, adolescent, and young adult cancer survivors: a report from the International Late Effects of Childhood Cancer Guideline Harmonization Group in collaboration with the PanCare Consortium. Lancet Oncol. 2019;20:e29–41. doi: 10.1016/S1470-2045(18)30858-1

30 Phelan M, Hayashi SS, Sauerburger K, et al. Prevalence of hearing screening failures in low-risk childhood cancer survivors. Pediatr Blood Cancer. 2022;69:e29437. doi: 10.1002/pbc.29437

31 Strebel S, Mader L, Jörger P, et al. Hearing loss after exposure to vincristine and platinum-based chemotherapy among childhood cancer survivors. EJC Paediatric Oncology. 2023;1:100017. doi: 10.1016/j.ejcped.2023.100017

32 Chen KS, Bach A, Shoup A, et al. Hearing loss and vestibular dysfunction among children with cancer after receiving aminoglycosides. Pediatr Blood Cancer. 2013;60:1772–7. doi: 10.1002/PBC.24631

33 Kompis M. Audiologie. 5th ed. Hogrefe AG 2022.

34 Panczak R, Berlin C, Voorpostel M, et al. The Swiss neighbourhood index of socioeconomic position: update and re-validation. Swiss Med Wkly. 2023;153:40028. doi: 10.57187/smw.2023.40028

35. Bundesamt für Statistik. Stadt/Land-Typologie 2020. 2024. https://www.atlas.bfs.admin.ch/maps/13/de/17847_17846_3191_227/27617.html (accessed 2 May 2025)

36 Harris PA, Taylor R, Minor BL, et al. The REDCap consortium: Building an international community of software platform partners. J Biomed Inform. 2019;95:103208. doi: 10.1016/J.JBI.2019.103208

37 Harris PA, Taylor R, Thielke R, et al. Research electronic data capture (REDCap)—A metadata-driven methodology and workflow process for providing translational research informatics support. J Biomed Inform. 2009;42:377–81. doi: 10.1016/J.JBI.2008.08.010

38 Brock PR, Knight KR, Freyer DR, et al. Platinum-induced ototoxicity in children: a consensus review on mechanisms, predisposition, and protection, including a new International Society of Pediatric Oncology Boston ototoxicity scale. J Clin Oncol. 2012;30:2408–17. doi: 10.1200/JCO.2011.39.1110

39 Moke DJ, Luo C, Millstein J, et al. Prevalence and risk factors for cisplatin-induced hearing loss in children, adolescents, and young adults: a multi-institutional North American cohort study. Lancet Child Adolesc Health. 2021;5:274–83. doi: 10.1016/S2352-4642(21)00020-1

40. Braun V, Clarke V. Thematic Analysis: A Practical Guide. SAGE 2021.

41 Jörger P, Nigg C, Schreck L, et al. Preprint: Hearing screening beyond the clinic: Childhood cancer survivors’ perceptions of a novel hearing screening program. medRxiv. 2025.

42 King N. Using Templates in the Thematic Analysis of Text. Essential Guide to Qualitative Methods in Organizational Research. 1 Oliver’s Yard, 55 City Road, London EC1Y 1SP United Kingdom : SAGE Publications Ltd 2004:256–70.

43 King N. Doing Template Analysis. Qualitative Organizational Research: Core Methods and Current Challenges. 1 Oliver’s Yard, 55 City Road London EC1Y 1SP : SAGE Publications, Inc. 2012:426–50.

44 Kuehni CE, Rueegg CS, Michel G, et al. Cohort Profile: The Swiss Childhood Cancer Survivor Study. Int J Epidemiol. 2012;41:1553–64. doi: 10.1093/ije/dyr142

45 Horowitz ME, Fordis M, Krause S, et al. Passport for care: implementing the survivorship care plan. J Oncol Pract. 2009;5:110–2. doi: 10.1200/JOP.0934405

46 Haupt R, Essiaf S, Dellacasa C, et al. The ‘Survivorship Passport’ for childhood cancer survivors. Eur J Cancer. 2018;102:69–81. doi: 10.1016/j.ejca.2018.07.006

47 Bexelius C, Honeth L, Ekman A, et al. Evaluation of an internet-based hearing test--comparison with established methods for detection of hearing loss. J Med Internet Res. 2008;10:e32. doi: 10.2196/jmir.1065

48 Rodríguez-Valiente A, Álvarez-Montero Ó, Górriz-Gil C, et al. Prevalence of presbycusis in an otologically normal population. Acta Otorrinolaringol Esp. 2020;71:175–80. doi: 10.1016/j.otorri.2019.05.002

49 Jörger P, Nigg C, Žarković M, et al. Awareness about the risk of hearing loss after ototoxic treatments in Swiss childhood cancer survivors. Patient Educ Couns. 2025;136:108764. doi: 10.1016/j.pec.2025.108764

50 Strebel S, Mader L, Sláma T, et al. Severity of hearing loss after platinum chemotherapy in childhood cancer survivors. Pediatr Blood Cancer. 2022;69:e29755. doi: 10.1002/pbc.29755

51. Bundesamt für Statistik. Wirtschaftliche Sozialhilfe - Gemeindeebene. 2021. https://www.bfs.admin.ch/bfs/en/home/statistics/social-security/social-assistance/recipients-social-benefits/monetary-social-assistance.assetdetail.23865240.html (accessed 15 August 2023)

52. Bundesamt für Statistik. Gemeindetypologie und Stadt/Land-Typologie 2012. 2017.

