## Supplementary Material for "Evaluation of a new, community-based screening program to detect hearing loss in adult childhood cancer survivors in Switzerland – Findings from the HEAR study"

### Supplementary Table 1: RE-AIM evaluation framework to evaluate the HEAR-study screening program.

| Definition of Domain^a^ | In the context of HEAR | Data sources |
| --- | --- | --- |
| Reach assesses number, proportion, and representativeness of individuals who are willing to participate, and reasons why or why not. | Assess the number and proportion of invited CCS who participated in the HEAR-study screening program and compare their clinical and sociodemographic characteristics to nonparticipants. | Childhood Cancer Registry (ChCR)  Participant baseline questionnaire |
| Effectiveness focuses on evaluating the program’s immediate and long-term impact (positive and negative), and whether the program achieves its objectives. | Assess the number and proportion of participants with (newly) detected hearing loss, the proportion of those who contacted a physician for further evaluation, and number of those acquiring a hearing aid after the screening program. | Audiogram results  Participant follow-up questionnaire 2 |
| Adoption assesses stakeholders’ willingness to adopt and implement the program. | Explore experiences and satisfaction with the screening program from perspectives of participants and the hearing aid shops | Participant interviews  Participant follow-up questionnaire 1 & 2  Hearing aid shop employee questionnaire and interviews |
| Implementation evaluates the feasibility of implementing the program within real-world contexts, assessing the extent to which the program was delivered consistently and as intended.  Maintenance investigates the possibility of continuing the program beyond the research project for long-term sustainability. | Evaluate if the program was implemented as intended and assess necessary requirements for program implementation from the perspectives of stakeholders involved. | Hearing aid shop employee questionnaire and interviews  Clinician group discussions |

^a^ Adapted according to definitions from Glasgow et al. 2019 [1].

### Supplementary Figure 1: CCS self-reported reasons for participation in the HEAR-study from baseline questionnaire. (N=404)

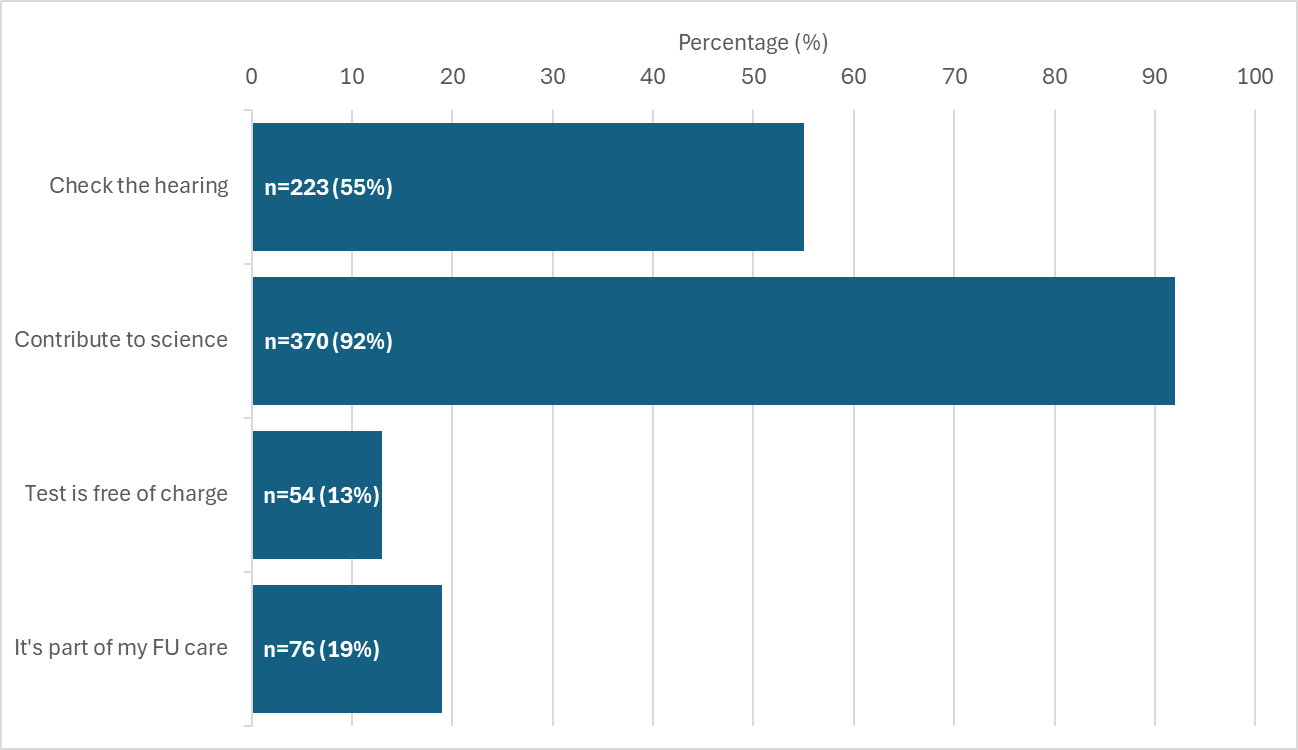

n=1 missing

Participants could select multiple answers, including “others”.

Abbreviations: n, number. FU, follow-up.

### Supplementary Table 2: Prevalence and severity of hearing loss according to SIOP-Boston ototoxicity scale, CPT-AMA, and self-reported hearing loss after the hearing test stratified by risk group (high or standard risk group based on treatment exposure).

| **Hearing loss** | **High risk group** | **Standard risk group** | **Total** |
| --- | --- | --- | --- |
|  | **n (%)** | **n (%)** | **n (%)** |
| **SIOP-Boston ototoxicity scale** | **N=83** | **N=236** | **N=319** |
| 0-1 | 46 (55%) | 202 (86%) | 248 (78%) |
| 2-4 | 37 (45%) | 34 (14%) | 71 (22%) |
| **CPT-AMA** | **N=83** | **N=236** | **N=319** |
| <20% | 71 (86%) | 230 (97%) | 301 (94%) |
| ≥20% | 12 (14%) | 6 (3%) | 18 (6%) |
| **Self-reported in follow-up questionnaire 2** | **N=65** | **N=193** | **N=258** |
| No impairment | 40 (62%) | 169 (88%) | 209 (81%) |
| Hearing impairment | 25 (38%) | 24 (12%) | 49 (19%) |

Abbreviations: SIOP, International Society of Pediatric Oncology. CPT-AMA, Council on Physical Therapy, American Medical Association.

Risk groups: High, treated with platinum agents (cisplatin/carboplatin) or cranial radiation ≥ 30 Gray. Standard, treated with any other chemotherapy or radiation to head, neck or spine with any dose.

Hearing loss results from the high risk group have been published previously [2].

CPT-AMA: The CPT-AMA scale is widely used in the Swiss health system to determine eligibility for hearing aid coverage by the old-age, survivors and disability insurance (OASI/IV) [3]. CPT-AMA results in percentage of hearing loss, calculated through weighted percentage for hearing threshold at frequencies between 500-4000 Hz [4]. We calculated the mean percentage of hearing loss of both ears and categorized it into not clinically relevant (<20%), or clinically relevant hearing loss (≥20%), based on the guidelines for ENT specialists on the clarification mandate for the reimbursement of hearing aids by the OASI and IV [3].

### Supplementary Table 3: General experiences of CCS participants with the hearing test.

|  | **SIOP 0-1 (N=232)** | **SIOP 2-4 (N=64)** | **Total^a^ (N=296)** | **P-value** |
| --- | --- | --- | --- | --- |
|  | n (%) | n (%) | n (%) |  |
| **Expectations for hearing test met^b^** |  |  |  | 0.04 |
| Yes | 210 (91%) | 51 (80%) | 261 (88%) |  |
| No | 1 (0%) | 1 (2%) | 2 (1%) |  |
| Partly | 20 (9%) | 12 (19%) | 32 (11%) |  |
| Missing | 1 (0.4%) | 0 (0%) | 1 (0.3%) |  |
| **Enough time for questions** |  |  |  | 0.61 |
| Yes | 223 (96%) | 63 (98%) | 286 (97%) |  |
| No | 9 (4%) | 1 (2%) | 10 (3%) |  |
| **Communication of results** |  |  |  | 0.18 |
| Oral and written | 177 (76%) | 46 (72%) | 223 (75%) |  |
| Oral | 49 (21%) | 13 (20%) | 62 (21%) |  |
| Written | 5 (2%) | 4 (5%) | 9 (3%) |  |
| Other | 1 (0%) | 1 (3%) | 2 (1%) |  |
| **Result was explained** |  |  |  | 0.66 |
| Yes, in detail | 206 (89%) | 55 (86%) | 261 (88%) |  |
| Yes, but I did not understand everything | 17 (7%) | 7 (11%) | 24 (8%) |  |
| No | 7 (3%) | 2 (3%) | 9 (3%) |  |
| Missing | 2 (0.9%) | 0 (0%) | 2 (0.7%) |  |
| **Information received on how to proceed** |  |  |  | 0.33 |
| Yes | 154 (66%) | 48 (75%) | 202 (68%) |  |
| No | 66 (28%) | 13 (20%) | 79 (27%) |  |
| Do not know | 12 (5%) | 2 (3%) | 14 (5%) |  |
| Missing | 0 (0%) | 1 (1.6%) | 1 (0.3%) |  |
| **Information was sufficient^c^** |  |  |  | 0.01 |
| Yes | 154 (66%) | 46 (72%) | 200 (68%) |  |
| No | 0 (0%) | 2 (3%) | 2 (1%) |  |
| Did not receive information on how to proceed or do not remember | 78 (34%) | 15 (23%) | 93 (31%) |  |
| Missing | 0 (0%) | 1 (1.6%) | 1 (0.3%) |  |
| **Would prefer discussing results directly with a doctor** |  |  |  | <0.01 |
| Yes | 40 (17%) | 26 (41%) | 66 (22%) |  |
| No | 192 (83%) | 38 (59%) | 230 (78%) |  |
| **Plans to discuss results with physician** |  |  |  | <0.01 |
| Yes | 4 (2%) | 14 (22%) | 18 (6%) |  |
| No | 211 (91%) | 39 (61%) | 250 (84%) |  |
| Do not know | 17 (7%) | 11 (17%) | 28 (9%) |  |
| **Travel duration to testing site** |  |  |  | 0.58 |
| <15min | 79 (34%) | 16 (25%) | 95 (32%) |  |
| 15-30min | 101 (44%) | 32 (50%) | 133 (45%) |  |
| 30-60min | 35 (15%) | 9 (14%) | 44 (15%) |  |
| >60min | 5 (2%) | 2 (3%) | 7 (2%) |  |
| Missing | 12 (5.2%) | 5 (7.8%) | 17 (5.7%) |  |
| **Duration of hearing test** |  |  |  | <0.05 |
| <30min | 141 (61%) | 26 (41%) | 167 (56%) |  |
| 30-60min | 91 (39%) | 37 (58%) | 128 (43%) |  |
| >60min | 0 (0%) | 1 (2%) | 1 (0%) |  |

Table with column percentages.

^a^ Participants completing follow-up questionnaire 1 (N=296)

^b^Further expectations:

- Wished more explanation about the diagram (audiogram)
- Wished for more information on hearing protection
- Wished for more information in general

^c^Open questions:

- Can I improve my hearing? Can I do something about it? (n=6)
- Should I check my hearing again? When? How often? (n=3)
- Were there anomalies? (n=2)
- What should I expect in the future? Will my hearing become worse? (n=2)
- How do I manage my hyperacusis?
- What is the reason for my hearing loss?

### Supplementary Figure 2: Answers from acousticians comparing study participants with usual customers. (Survey, N=13).

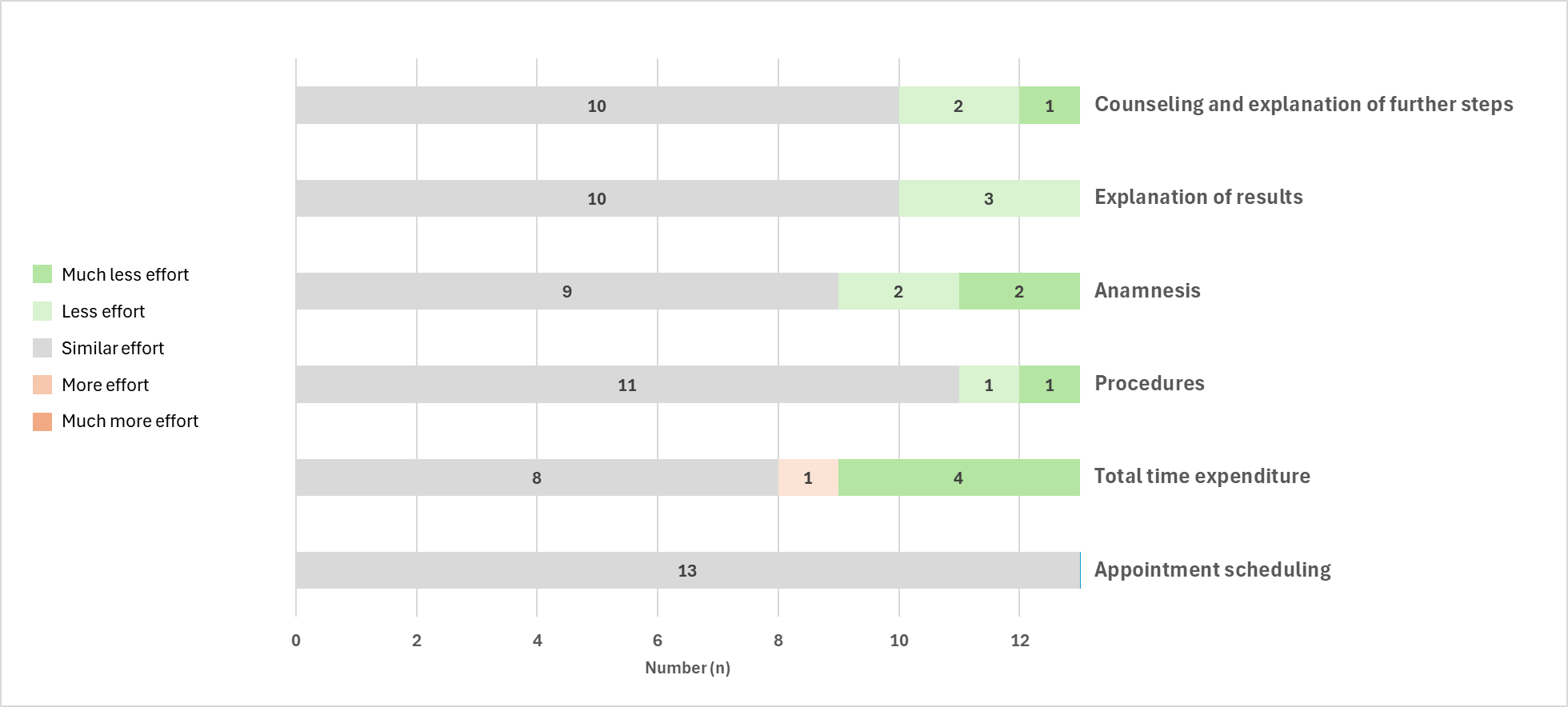

Abbreviations: n, number.

### Supplementary Figure 3: Potential barriers as perceived by acousticians completing a survey (N=13).

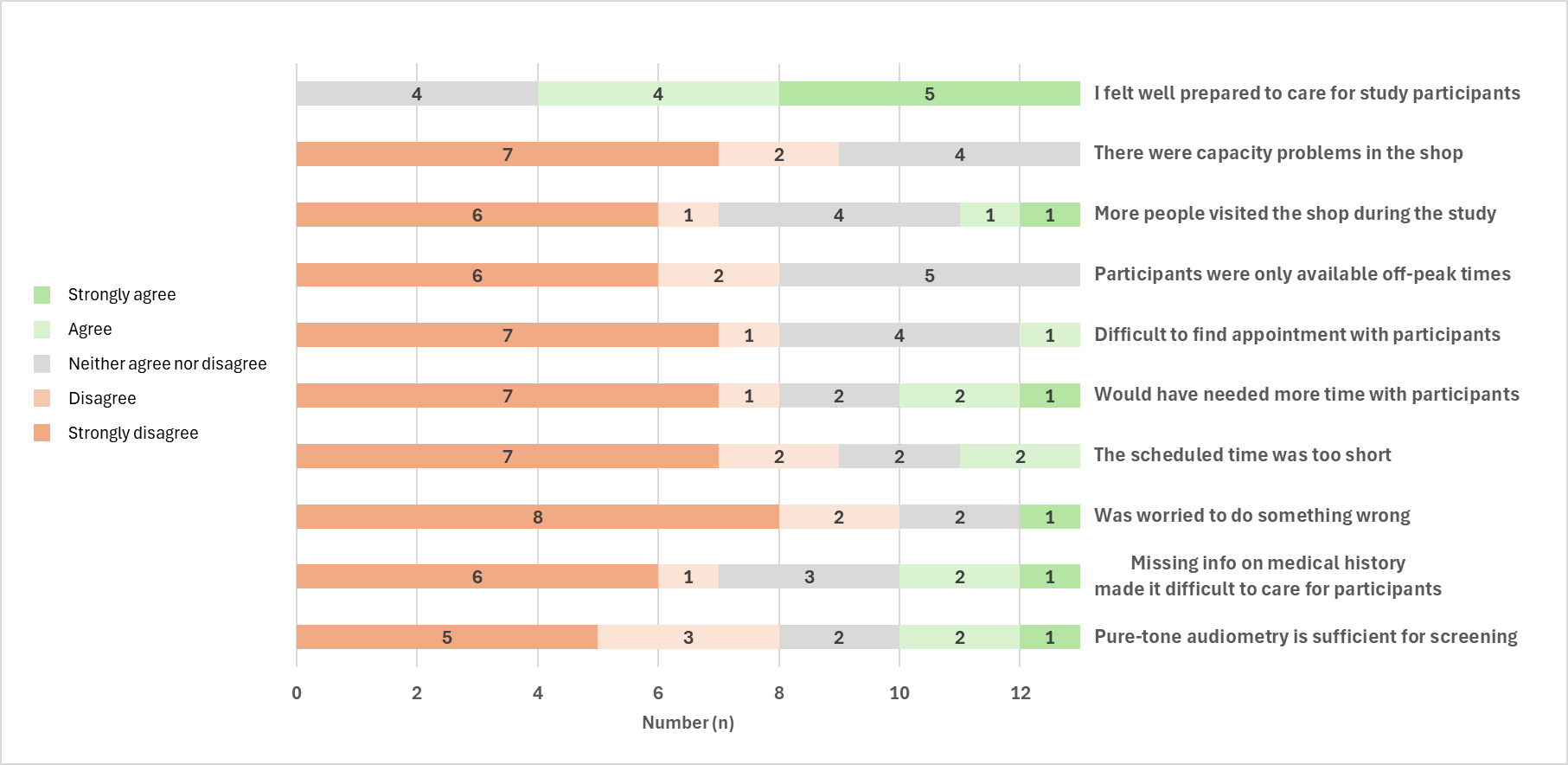

Abbreviations: n, number.

References

1	Glasgow RE, Harden SM, Gaglio B, *et al.* RE-AIM Planning and Evaluation Framework: Adapting to New Science and Practice With a 20-Year Review. *Front Public Health*. 2019;7:64. doi: 10.3389/fpubh.2019.00064

2	Jörger P, Nigg C, Žarković M, *et al.* Awareness about the risk of hearing loss after ototoxic treatments in Swiss childhood cancer survivors. *Patient Educ Couns*. 2025;136:108764. doi: 10.1016/j.pec.2025.108764

3	Bundesamt fuer Sozialversicherungen BSV. Richtlinien fuer ORL- Expertenaerzte zum Abklaerungsauftrag zur Verguetung von Hoergeraeten durch die Sozialversicherungen IV und AHV. 2018.

4	Council on Physical Therapy American Medical Association. TENTATIVE STANDARD PROCEDURE FOR EVALUATING PERCENTAGE OF USEFUL HEARING LOSS IN MEDICOLEGAL CASES. *Archives of Otolaryngology - Head and Neck Surgery*. 1942;36:590–2. doi: 10.1001/archotol.1942.03760040144015
